# Development and Scoring of a Diet Quality Screener for Hispanic/Latino Adults Using National Health and Nutrition Examination Survey Data: The ¿SABE Lo Que Come? Tool

**DOI:** 10.64898/2026.09.09.26362652

**Authors:** Margaret S. Pichardo, Vasu Shandar, Ryan Quinn, Victoria McReynolds, Kate Townsend Creasy, Gary D. Wu, Charlene Compher

## Abstract

**Background:** Few dietary screeners are available for use with Hispanic/Latino adults in clinical or research settings, despite this group having a disproportionate burden of diet-related chronic disease risk amenable to nutrition intervention and counseling. We previously developed the 36-item Penn Healthy Diet (PHD) screener, which correlates with the Healthy Eating Index (HEI), and adapted it culturally and linguistically for use among Hispanic/Latino adults as the Screener of Alimentary Behaviors and Eating (SABE).

**Objective:** We aimed to develop a scoring algorithm for the SABE screener using data from Hispanic/Latino adults in the National Health and Nutrition Examination Survey (NHANES).

**Methods:** We used NHANES 2017–2018 dietary recall data from 1,126 Hispanic/Latino adults to simulate responses to SABE screener items. Using Spearman rank correlations, we examined the association between simulated SABE items and HEI-2020 components and validation using the Alternative Healthy Eating Index (AHEI)-2010. Items achieving |ρ| ≥ 0.3 with any HEI-2020 component were retained in a simple scoring algorithm. A regression-weighted algorithm was then derived using ordinary least squares regression of all screener items on HEI-2020 total score. Reproducibility was evaluated in an independent validation sample of 1,633 Hispanic/Latino adults from NHANES cycle 2015–2016.

**Results:** Twelve of 24 screener items were moderately or strongly associated with HEI-2020 components, forming a simple scoring algorithm with a range of 0–60 points. The simulated SABE score was strongly correlated with HEI-2020 (ρ = 0.73-0.74) and moderately correlated with AHEI-2010 (ρ = 0.64-0.67). The regression-weighted algorithm improved HEI-2020 correlations (ρ = 0.80) but not for AHEI-2010 (ρ = 0.62-0.65).

**Conclusions:** The SABE screener is a brief and feasible dietary screener with demonstrated strong validity and reproducibility for assessing diet quality in Hispanic/Latino adults. Further validation of this screener is needed in Hispanic/Latinos subgroups of diverse heritage and acculturation level, and in real-world populations.

## INTRODUCTION

There is increasing recognition of the importance of dietary intake patterns in the prevention and management of chronic diseases (1, 2). Dietary data collection is crucial for providing personalized, evidence-based nutritional guidance to patients. Despite the demonstrated clinical benefits of even brief dietary counseling (3), dietary screening and counseling remain largely outside the scope of routine medical visits (1). Key barriers to widespread implementation in primary care include lack of training, perceived futility, and inadequate reimbursement (1). Further, traditional dietary assessment tools impose additional time and resource burden on providers (4), while participants frequently cite lack of personal time or difficulties accessing online platforms as reasons for non-completion (5). In research settings, dietary data collection is costly and labor-intensive, often requiring trained interviewers and substantial data entry and processing (4, 6). Even among engaged participants, systematic and random measurement errors are common (7), with misreported information occurring in approximately 25% of participants (8), particularly among individuals with obesity (9). Additionally, many of these dietary assessments require computer literacy and internet access, further limiting their utility for assessing dietary quality.

Dietary screening tools have been developed for specific U.S. Hispanic/Latino subgroups (10–13), but brief screeners with broader generalizability to adults of diverse Hispanic/Latino heritage are needed. These screeners serve as a practical solution for the logistical constraints of busy clinical and research settings and demonstrate concordance with gold-standard 24-hour dietary recalls (14). Targeted dietary screeners for Hispanic/Latino patients are uniquely necessary, as Hispanic/Latino research participants experience additional barriers related to time, trust, and cultural relevance when participating in nutrition research, contributing to reduced enrollment and high attrition rates (15, 16). Furthermore, when adapting dietary screeners to Hispanic/Latino adults, scoring algorithms derived from general adult samples of predominantly non-Hispanic White backgrounds may not accurately characterize dietary quality in Hispanic/Latino adults. Indeed, studies consistently show lower validity of food frequency questionnaires among Hispanic/Latino adults relative to non-Hispanic White adults, even with adaptation, likely introducing error and bias into dietary estimates (17). These findings underscore the need for population-specific calibration of dietary screening tools and scoring algorithms for use among Hispanic/Latino adults.

To address this need, our group previously developed the Penn Healthy Diet (PHD), a novel dietary screener for use in a community and clinical setting (18). PHD is a 36-item dietary screener that is moderately correlated with Healthy Eating Index (HEI)-2015 (Spearman rho 0.75) (18) and has been widely accepted for use by patients and dietitians (19). This tool was subsequently culturally and linguistically adapted for use in Spanish-speaking Hispanic/Latino adults as the, *¿SABE Lo Que Come?* (SABE) screener. Via content validation of the instrument for use in a general adult population that self-identified as Hispanic/Latino, the SABE screener was found both clear (item-level content validity index (I-CVI): 0.865) and relevant (I-CVI: 0.969) (20).

In this subsequent study, we aim to develop a scoring algorithm for the SABE screener using National Health and Nutrition Examination Survey (NHANES) data for Hispanic/Latino adults. Our specific objectives are to [1] compute dietary quality indices (HEI and Alternative Healthy Eating Index (AHEI) scores) using dietary recall data from Hispanic/Latino adults; [2] compute *simulated responses* to the SABE screener items using recall data; [3] compare simulated screener responses to HEI and AHEI components computed from recalls; and [4] create a simple and regression weighted scoring algorithm for screener responses and correlate with dietary quality indices.

## METHODS

### Data Source and Study Population

NHANES is a nationally representative, cross-sectional survey of the non-institutionalized U.S. civilian population conducted by the National Center for Health Statistics (21). We extracted data from the 2017– 2018 and 2015–2016 cycles. The survey employs a complex, multistage probability sampling design with oversampling of Hispanic persons, non-Hispanic Black persons, non-Hispanic Asian persons, older adults, and low-income individuals (22).

The analytic sample was restricted to self-identified Hispanic/Latino adults aged 18 years and older, defined as participants with NHANES race/ethnicity codes of Mexican American or Other Hispanic. Participants were subsequently excluded if they had nonpositive dietary recall sample weights, missing total energy intake, or zero HEI-2020 total scores, the latter indicating missing or invalid dietary recall data (see **Figure 1** for a flow diagram of study exclusions). No additional exclusions were made for extreme dietary values given that excluding participants based on caloric thresholds would risk introducing selection bias in a population-based sample (23). Observed extreme values (minimum 14 kcal, maximum 11,710 kcal) were retained in the analytic samples. The final analytical sample was 1,126 participants in the 2017-2018 cycle (training dataset) and 1,633 participants in the 2015–2016 cycle (validation dataset).

**Figure 1.**
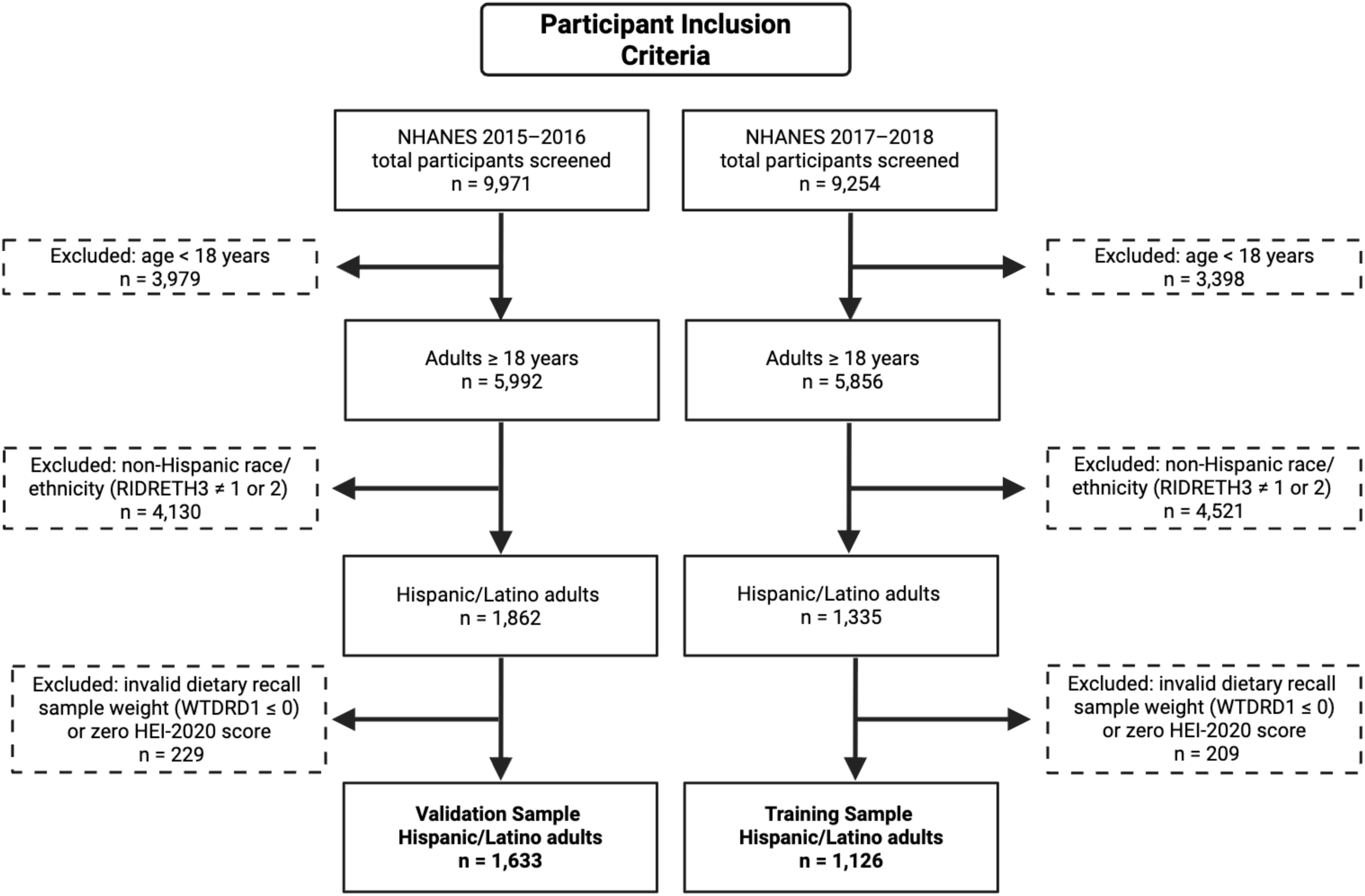
Flow diagram for participant inclusion showing sequential exclusions from the NHANES 2017–2018 and 2015–2016 cycles that yielded the training (n = 1,126) and validation (n = 1,633) samples of Hispanic/Latino adults.

**Figure 2.**
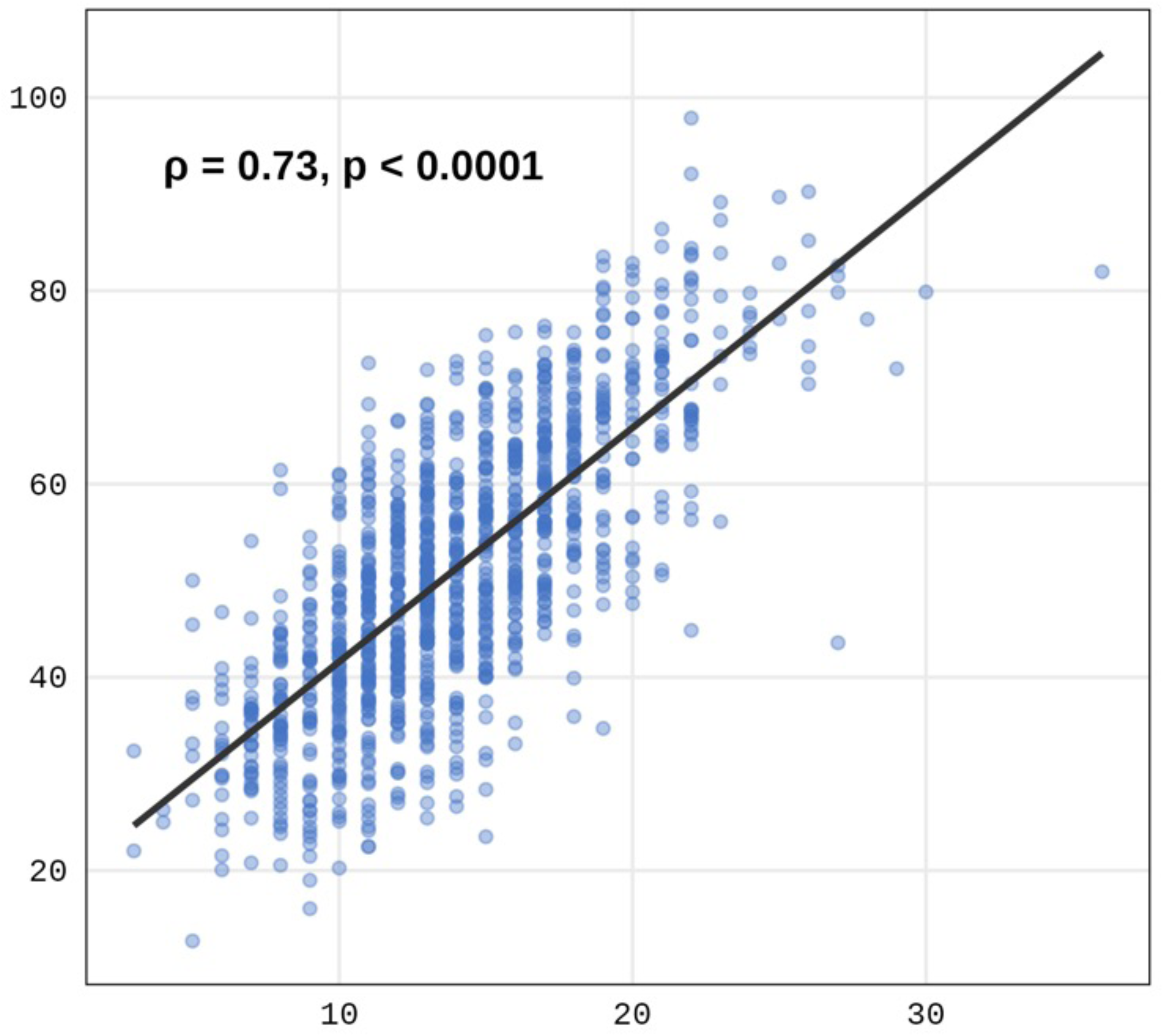
Scatter plot and regression line comparing the simulated SABE screener score (x-axis) and the Healthy Eating Index (HEI)-2020 score (y-axis) among Hispanic/Latino adults in NHANES 2017–2018 (N = 1,126).

### Dietary Assessment

Dietary intake was assessed using one 24-hour dietary recall administered in person at the NHANES Mobile Examination Center by trained interviewers using the USDA Automated Multiple-Pass Method (24). Food group intake was quantified using the Food Patterns Equivalents Database (FPED), which converts individual food items from the dietary recall into standardized food group servings (e.g., cup-equivalents of fruit, ounce-equivalents of whole grains) (25). Nutrient-level intake data (total energy, fatty acids, sodium, added sugars) were obtained from the NHANES Total Nutrient Intakes file.

### Simulated Screener Responses

Following the methodology used in scoring the PHD screener (18), SABE screener item responses were simulated from the NHANES dietary recall data from Hispanic/Latino participants. For each of 18 food group variables, continuous FPED intake values were organized into responses on a 0–5 scale representing frequency of intake: 0 servings/day = 0; 0 to <1 serving/day = 1; ≥1 to <2 servings/day = 2; ≥2 to <3 servings/day = 3; ≥3 to <4 servings/day = 4; ≥4 servings/day = 5.

Screener items derived from a single corresponding FPED (or NHANES dietary behavior) variable were categorized directly into the 0–5 point screener scale without unit conversion or variable combination. These included fruit juice, other/whole fruit, dark green vegetables, whole grains, refined grains, milk, yogurt, cheese, eggs, nuts/seeds, and sweetened beverages. Protein-based food groups (meat, cured meat, poultry) were first converted from ounce-equivalents to 3-ounce servings before categorization. Combined screener variables were created for seafood (high-omega-3 + low-omega-3 seafood), plant protein (soy + legumes), red/orange vegetables (red/orange vegetables + tomatoes) and desserts (desserts + bakery items) by summing component FPED variables. Six yes/no behavioral items were also created from FPED food category variables (sugar/honey in coffee, artificial sweetener use, cream in coffee, full-fat dairy products, butter/gravy use, and olive/vegetable oil use). Of note, these behavioral variables do not have equivalent subcomponents in gold standard dietary quality indices like HEI or AHEI.

It is important to note that because both the simulated screener responses and the dietary quality index scores are derived from the same 24-hour dietary recall data, the resulting correlations represent the theoretical ceiling of screener performance. In clinical practice, when patients self-report dietary intake from memory, measurement error as well as differences in the estimation of portion sizes by patients compared to trained interviewers or differences in food categories may introduce errors that reduce correlations. The PHD validation study demonstrated this directly: simulated correlations with HEI were ρ = 0.75, while correlations using actual patient-reported screener responses were ρ = 0.59 (26). The present analysis establishes the upper bound of SABE screener performance in the Hispanic subgroup; subsequent validation with patient-administered screener data would be needed to estimate real-world performance.

### Dietary Quality Index Calculation

The primary dietary quality index used to evaluate the screener was the HEI-2020. HEI-2020 total and component scores were pre-calculated and provided in the analytic dataset using the National Cancer Institute scoring algorithm. The HEI-2020 measures adherence to the 2020–2025 Dietary Guidelines for Americans and includes 13 components, with a total score range of 0–100. Higher scores indicate greater adherence to dietary guidelines (27).

In sensitivity analysis, the AHEI-2010 was used to validate the screener. AHEI-2010 scores were computed following the scoring criteria published by Chiuve et al. (28). Unlike the HEI, which measures dietary guideline adherence, the AHEI was designed based on foods and nutrients predictive of chronic disease risk, particularly cardiovascular disease and diabetes (28). Ten of the 11 AHEI components were scored on a 0–10 scale using linear interpolation between minimum (0 points) and maximum (10 points) cut points: vegetables excluding potatoes and including tomatoes (0–5 servings/day), whole fruit (0–4 servings/day), whole grains (0–75 g/day for women, 0–90 g/day for men), sugar-sweetened beverages and fruit juice (≥1–0 servings/day, reverse scored), nuts and legumes (0–1 serving/day), red and processed meat (≥1.5–0 servings/day, reverse scored), long-chain omega-3 fatty acids (0–250 mg/day), polyunsaturated fatty acids excluding eicosapentaenoic acid (EPA) and docosahexaenoic acid (DHA) (2–10% of energy), sodium (scored by decile of distribution), and alcohol (gender-specific moderate intake scoring with nondrinkers receiving 2.5 points). The trans-fat component was omitted because NHANES 2017–2018 does not report trans fatty acid intake separately. Total AHEI-2010 scores ranged from 0 to 100, with higher scores indicating a dietary pattern more strongly associated with reduced chronic disease risk.

### Statistical Analysis

Descriptive statistics for continuous variables were computed as weighted means and standard errors, accounting for the complex survey design through specification of strata, primary sampling units, and dietary recall sample weights. Survey weights were calculated in the entire NHANES cohort, prior to the application of race and ethnicity exclusions. Categorical variables were summarized as weighted percentages and counts. Food group intake was compared across Hispanic subgroups (Mexican American, Other Hispanic) and Non-Hispanic adults using survey-weighted means.

A two-step approach was employed to develop and validate the SABE scoring algorithm, consistent with the PHD scoring approach (18). First, Spearman rank-order correlations were computed between each of the 24 simulated screener item responses (18 continuous + 6 yes/no), all 13 individual HEI-2020 component scores, and total HEI-2020 scores. Items were retained in the SABE scoring algorithm if they achieved a moderate or strong correlation (|ρ| ≥ 0.3) with any individual HEI-2020 component. Second, the retained items were summed to compute a total SABE screener score, and this single composite score was correlated with the total HEI-2020 score. This two-step approach was also applied to AHEI-2010 to evaluate validity between the two indices.

To determine whether differential weighting of food groups could improve predictive accuracy, ordinary least squares regression was used to model HEI-2020 total score as a function of all 24 (18 continuous and 6 yes/no) screener items simultaneously. The resulting unstandardized beta coefficients represent the independent contribution of each food group to predicting overall diet quality in Hispanic adults. The regression-weighted SABE score was computed by multiplying each screener item response by its beta coefficient and adding the model intercept, placing the score on an approximately 100-point scale. Ordinary least squares regression models estimated beta coefficients and 95% confidence intervals (CI).

All scoring algorithms (simple and regression-weighted) developed in the 2017–2018 NHANES cycle were applied without modification to the independent 2015–2016 NHANES cycle. All statistical tests were two-sided, with significance set at α = 0.05. All analyses were conducted using SAS Viya Workbench Version 2026.04 (SAS Institute, Cary, NC) (29). Heatmap visualizations of correlation matrices were generated using the ggplot2 package in R (30) via Google Collaboratory (31). This study follows the STROBE-nut (Strengthening the Reporting of Observational Studies in Epidemiology–Nutritional Epidemiology) guidelines (32).

## RESULT

### Descriptive characteristics

**Figure 1** depicts a flow diagram for participant inclusion, showing sequential exclusions from the NHANES 2017–2018 and 2015–2016 cycles that yielded the training (n = 1,126) and validation (n = 1,633) samples of Hispanic/Latino adults.

**Table 1** shows descriptive characteristics of 1,126 Hispanic/Latino adult participants in NHANES 2017-2018. The mean (± standard error) age was 41 (± 0.87) years, with 51% identifying as female, 57% self-reporting Mexican American heritage, and approximately 85% reporting less than a college education. The mean (± SE) body mass index (BMI) was 30.38 (± 0.30). **Supplemental Table 1** shows the percent intake of HEI and AHEI score components for Mexican American, Other Hispanic and Non-Hispanic adults in NHANES cycles 2017-2018.

**Table 1:** Descriptive statistics for Hispanic/Latino adults in NHANES cycle 2017-2018. Values are weighted means ± standard error. Weights account for the complex survey design of NHANES (WTDRD1).

| Characteristic | Number | Mean | Std Error of Mean | 95% CL for Mean |  |
| --- | --- | --- | --- | --- | --- |
| Age (years at study entry) | 1,126 | 41.45 | 0.87 | 39.59 | 43.32 |
| Income (ratio of family income to poverty) | 1,057 | 2.34 | 0.07 | 2.18 | 2.49 |
| Body Mass Index (kg/m²) | 1,126 | 30.38 | 0.30 | 29.75 | 31.02 |
| Daily caloric intake (kcal) | 1,126 | 2,183 | 48.75 |  |  |
| Healthy Eating Index (HEI)-2020 |  | 50.79 | 0.76 |  |  |
| Alternative HEI |  | 38.40 | 0.82 |  |  |
| Sex (n= 1,126) |  | Weighted Frequency |  | Row Percent |  |
| Male | 532 | 19,762,007 |  | 49.32 |  |
| Female | 594 | 20,303,223 |  | 50.68 |  |
| Self-reported Hispanic subgroup (n= 1,126) |  |  |  |  |  |
| Mexican American | 673 | 22,913,246 |  | 57.19 |  |
| Other Hispanic | 453 | 17,151,985 |  | 42.81 |  |
| Education level (n = 1,057) |  |  |  |  |  |
| Less than 9th grade | 265 | 5,701,148 |  | 15.12 |  |
| 9th–11th grade | 171 | 4,216,475 |  | 11.18 |  |
| High school graduate/GED | 221 | 11,299,687 |  | 29.96 |  |
| Some college or AA degree | 277 | 10,998,455 |  | 29.16 |  |
| College graduate or above | 120 | 5,472,577 |  | 14.51 |  |

### Simple scoring algorithm

We examined correlations between individual screener items and HEI components (**Figure 3**) and identified food groups with moderate or strong positive correlation across both NHANES cycles, including fruit juice, whole fruit, dark green vegetables, red/orange vegetables, whole grains, milk, nut/seeds, seafood, and plant proteins. Certain screener items were negatively correlated with HEI subcomponents recommended in moderation. For example, sweetened beverage on the screener was negatively correlated with the added sugar subcomponent, cheese on the screener was negatively correlated with the saturated fats subcomponent, and refined grain was negatively correlated with the refined grain subcomponent. Screener items that moderately or strongly positively correlated with the AHEI-2010 components included whole fruit, whole grain, nuts/seeds, and plant proteins, while sweetened beverages correlated negatively. Overall, the screener successfully identified key food groups associated with dietary quality.

**Figure 3.**
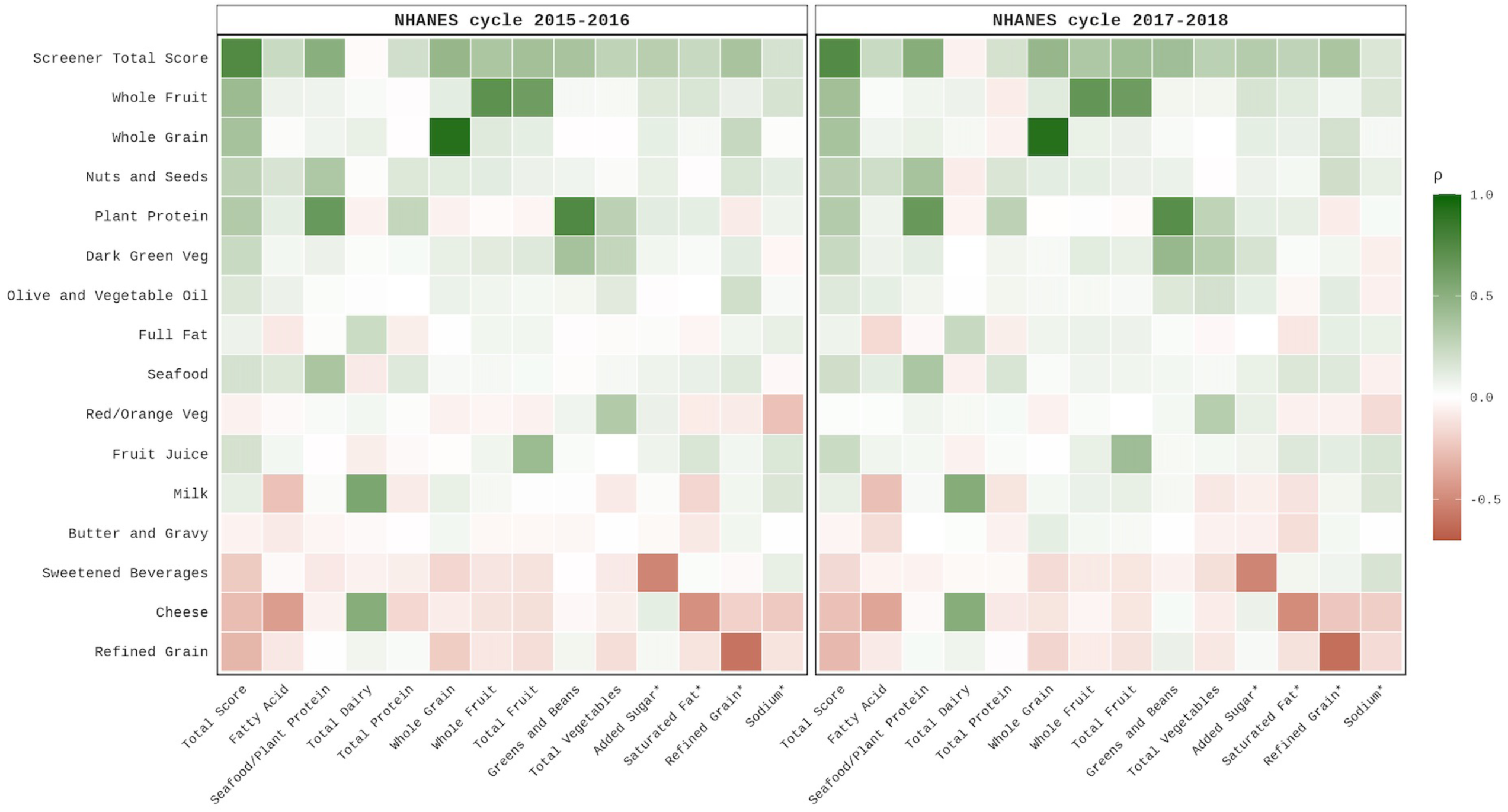
Heat map of rank-ordered Spearman correlations between individual simulated SABE screener items (y-axis) and HEI-2020 total and subcomponents (x-axis). Correlations colored green are positively associated and red are negatively associated. *Denotes HEI-2020 components with recommended consumption in moderation.

**Figure 4.**
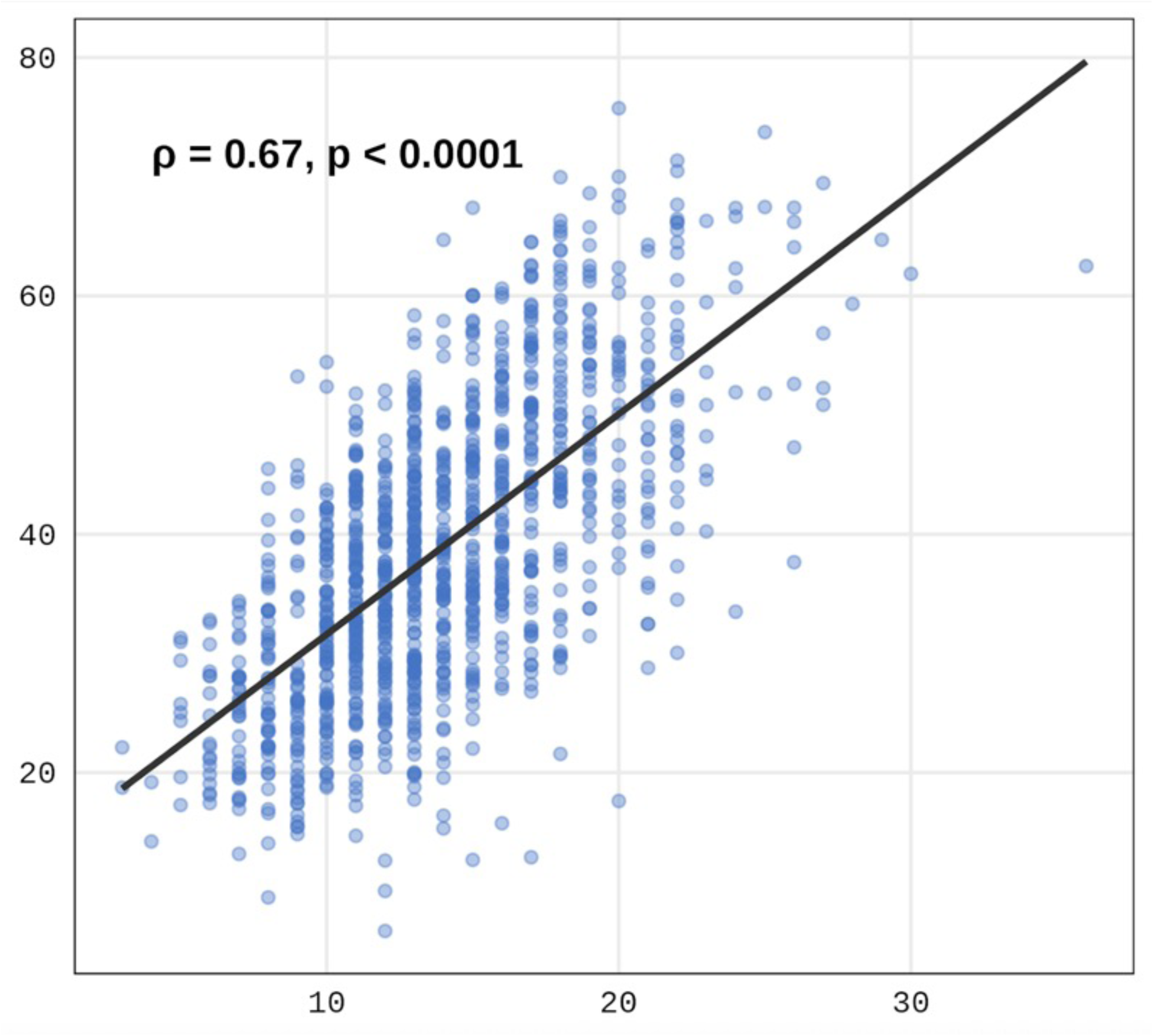
Scatter plot and regression line comparing the simulated SABE screener score (x-axis) and total AHEI-2010 score (y-axis) score among Hispanic/Latino adults in NHANES 2017–2018 (N = 1,126).

**Figure 5.**
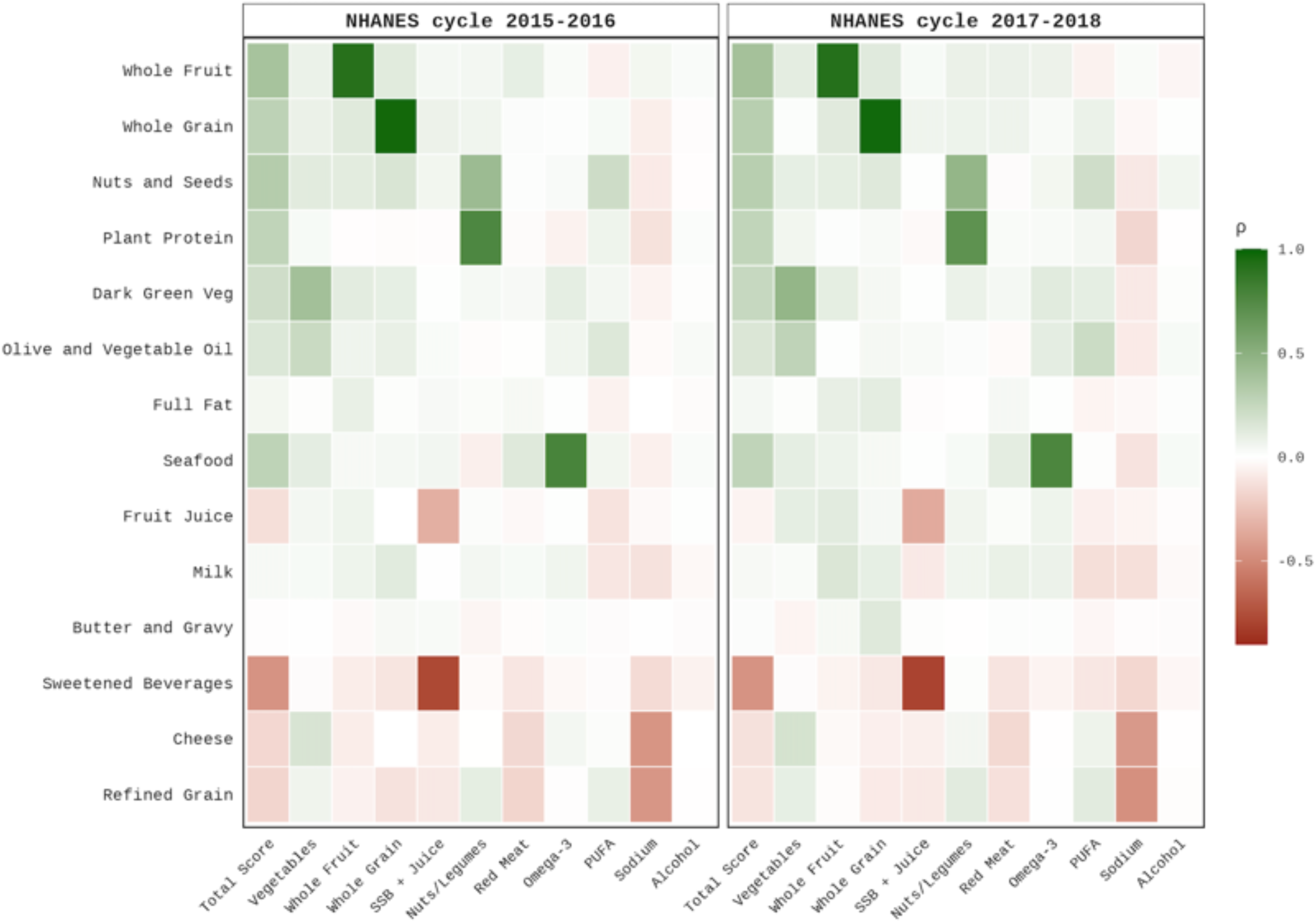
Heat map of rank-ordered Spearman correlations between individual simulated SABE screener items (y-axis) and AHEI-2010 total and subcomponents (x-axis). Correlations colored green are positively and red are negatively associated.

To generate a scoring algorithm, we retained screener items strongly or moderately positively associated with HEI-2020 components (|ρ| ≥ 0.3) and assigned points based on the number of servings reported in the predicted screener response (**Supplemental Table 2**). The SABE screener showed weak correlations with HEI-2020 components for olive/vegetable oil, full-fat dairy and butter/gravy, which did not meet our threshold for inclusion. The SABE screener was consistent with the PHD screener in excluding yogurt, eggs, poultry, red meat, cured meat, and desserts.

The SABE simple scoring algorithm sums the screener responses for retained items. Nine items with positive correlations were scored in the forward direction (0–5 points each): fruit juice, whole fruit, dark green vegetables, whole grains, milk, nuts/seeds, seafood, and plant protein. Three items with negative correlations were reverse scored: refined grains, cheese, and sweetened beverages. The reverse score was computed as 15 (sum of reverse items), yielding 0–5 points each when intake is lowest to highest, respectively. The total SABE simple score potentially ranges from 0 to 60, with higher scores indicating a healthier diet.

Three items included in the original PHD simple scoring algorithm were excluded from the SABE algorithm because they did not meet the |ρ| ≥ 0.3 threshold in the Hispanic subgroup: all three yes/no behavioral items: olive/vegetable oil use (ρ = 0.19), full-fat dairy use (ρ = 0.24), and butter/gravy use (ρ = −0.15) (18). Consequently, the potential range of the SABE score (0–60) is narrower than that of the PHD score (0–63).

### Regression weights predicting reference indices

In a multivariable model using all 24 screener items (18 continuous frequency items and 6 yes/no behavioral items) to predict total HEI-2020 score, the full item set explained 65% of the variance in diet quality (R^2^ = 0.649; **Supplemental Table 3**). The items which demonstrated the strongest positive associations were plant protein (standardized β = 0.35), whole fruit (β = 0.27), and whole grains (β = 0.24), while refined grains (β = −0.24) and cheese (β = −0.16) demonstrated the strongest negative associations. Several items not currently in the simple algorithm such as, desserts (β = −0.13), yogurt (β = 0.09), red meat (β = −0.09), and cured meat (β = −0.08), contributed independently at magnitudes comparable to retained items, whereas poultry, and three yes/no behavioral items (sugar/honey, artificial sweetener, and creamer) were nonsignificant (*P* = 0.10).

A multivariable regression model was also produced for the AHEI-2010 (R² = 0.714; **Supplemental Table 4**). Consistent with the HEI-2020 model, whole fruit (standardized β = 0.27), plant protein (0.25), and nuts/seeds (β = 0.22) demonstrated the strongest positive associations. However, sweetened beverages (β = −0.33), red meat (β = −0.27), and cured meat (β = −0.23) demonstrated the strongest negative associations, each demonstrating a stronger association than in the HEI-2020 model. Notably, fruit juice was negatively associated with the AHEI-2010 (β = −0.11) despite being positively associated with the HEI-2020 (β = 0.15). Additionally, refined grains did not significantly predict the AHEI-2010 (β = −0.02, *P* = 0.18) despite demonstrating moderate association with the HEI-2020.

### Screener performance against reference indices

Among 1,126 Hispanic/Latino participants in NHANES cycle 2017-2018, the mean (± SE) HEI-2020 score was 51.03 (± 13.93) and mean total daily caloric intake was 2,183.4 (± 48.75). The simulated SABE screener mean (± SE) created from the simple algorithm was 14.79 (± 4.38). This SABE total score was strongly associated with the HEI-2020 total score (Spearman ρ = 0.73; *P* < 0.0001; **Figure 2**; **Table 2**), with separate validation in NHANES 2015-2016 cycle (Spearman ρ = 0.75; *P* < 0.0001; **Table 2**). The mean (± SE) AHEI-2010 total score was 38.79 (± 12.31). The SABE total score was moderately associated with the AHEI-2010 total score (Spearman ρ = 0.67; P < 0.0001; **Supplemental Table 1**) with strong individual associations in the whole fruit (ρ = 0.99) and whole grains (ρ = 0.99) screener items (**Supplemental Table 5**). Estimates were consistent across the two independent NHANES cycles supporting the reproducibility of the scoring algorithms.

**Table 2.** Spearman rank correlations (ρ) between the simulated SABE screener total score and dietary quality indices in Hispanic/Latino adults across two NHANES cycles.

| Dietary Index | Simple Algorithm |  | Regression-Weighted |  |
| --- | --- | --- | --- | --- |
|  | 2017–18 | 2015–16<br>Validation | 2017–18 | 2015–16<br>Validation |
| HEI-2020 | 0.73 | 0.74 | 0.80 | 0.80 |
| AHEI-2010 | 0.67 | 0.64 | 0.65 | 0.62 |
| Notes: The simple algorithm is the sum of 9 forward-scored and 3 reverse-scored food group items (score range 0–60). The regression-weighted algorithm has screener items weighted by beta coefficients derived from ordinary least squares regression predicting HEI-2020 total score in the 2017–2018 development sample, applied without modification to the 2015–2016 validation sample. |  |  |  |  |

The regression weighted SABE algorithm showed comparable or stronger correlations with reference indices relative to the simple algorithm. In the 2017–2018 development sample, the regression-weighted score was associated with HEI-2020 total score (Spearman ρ = 0.80; P < 0.0001; **Table 2**) and moderately associated with AHEI-2010 total score (Spearman ρ = 0.65; P < 0.0001; **Table 2**). In the independent 2015– 2016 validation sample, the regression-weighted algorithm remained strongly associated with HEI-2020 (Spearman ρ = 0.80; P < 0.0001; **Table 2**) and moderately associated with AHEI-2010 (Spearman ρ = 0.62; P < 0.0001; **Table 2**). The regression-weighted approach improved HEI-2020 correlation in the validation sample relative to the simple algorithm, while AHEI-2010 correlations were similar between the two scoring approaches.

### DISCUSSION (4 pages max, double spaced)

¿SABE Lo Que Come? (SABE) is a brief dietary screener for Spanish-speaking Hispanic/Latino adults, developed for use in clinical, community, and research settings (20). Using NHANES dietary recall data, we found that simulated SABE responses were strongly correlated with HEI-2020 (ρ = 0.73) and moderately correlated with AHE-2010 scores (ρ = 0.67), supporting SABE as a rapid tool for quantifying diet quality and informing dietary counseling in this population. SABE was adapted from an English-language screener validated against HEI-2015 and content validated among a bilingual sample of health professionals and lay participants, a process that captured regional variation in Spanish dialects and supports its use across diverse Hispanic/Latino heritage groups (20).

One key difference between the scoring of SABE and the original Penn Healthy Diet (PHD) screener is the exclusion of yes/no items for olive/vegetable oil, full-fat dairy and butter/gravy, likely reflecting low reported intake of these items in NHANES as only 6.2% of participants reported any intake in the FPED butter, animal fat, and gravy categories, which captures fats explicitly added during preparation but not fats embedded within dishes, such as lard (*manteca*) used in cooking tamales or refried beans. While Mexican Americans report more frequent use of oil than lard or butter (33), many traditional dishes are still prepared with lard (11) and this use may go unrecorded unless recall methodology specifically probes for cooking ingredients. This likely reflects a limitation of NHANES recall methodology for culturally specific cooking practices rather than a true absence of lard use, though traditional, less-acculturated Mexican diets are also associated with lower overall fat intake (34). Future SABE iterations will add a culturally adapted item probing for lard use directly.

Fruit juice scoring also diverged between indices, where the HEI-2020 scores juice forward as equivalent to whole fruit (ρ = 0.45 with the Total Fruit component, β = +2.55), while AHE-2010 reverse-scores juice within the sugar-sweetened beverage category given its cardiovascular risk potential (ρ = −0.39, β = −1.67) (28). Because fruit juice and fruit-flavored beverages such as *agua fresca* are common in Hispanic/Latino diets, SABE’s current forward-scoring of juice may overstate diet quality for high consumers. This divergence likely also explains why SABE’s simulated AHEI-2010 correlation was weaker than its HEI- 2020 correlation, and lower than AHEI scores reported in real-world Hispanic/Latino populations, consistent with evidence that AHEI’s performance varies by heritage group and does not uniformly capture Hispanic/Latino dietary patterns (35). Future SABE iterations will reclassify fruit juice with sugar-sweetened beverages and restrict the whole-fruit item to exclude juice.

A further methodological limitation involves the classification of tortillas. FPED categorizes corn tortillas as whole grain, but SABE groups all tortillas with white bread, pasta, and rice as refined grains. Because self-administered responses will likely reflect this grouping regardless of tortilla type, real-world administration may inflate real-world correlations and understate whole-grain correlations, particularly for Mexican-heritage populations with high corn tortilla consumption. Future SABE versions will distinguish corn and flour tortillas and tortilla chips, to better align with FPED definitions. Separately, whole grain intake showed near-perfect correlation with the HEI whole grain subcomponent, largely because more than half of the sample reported no whole grain intake. This reflects a zero-inflation pattern also seen broadly across NHANES samples, not unique to Hispanic/Latino adults (36), which produces a large tied-rank group rather than reflecting true measurement precision.

Culturally and linguistically adapted tools like SABE are likely to improve the validity, feasibility and acceptability of dietary assessment among Hispanic/Latino adults (37, 38). Such adaptation may also improve engagement with nutrition counseling and research via increased language concordance that facilitates trust and satisfaction with the care received (39), while cultural tailoring drives greater behavioral engagement with the screener itself (40), including increased comprehension, acceptability, and completion rates (41).

Other Hispanic/Latino-specific dietary screeners exist for Mexican American (10, 11, 13), Puerto Rican (12), and Dominican (12) populations, but SABE was designed for broader applicability. It was not, however, built to capture every culturally specific food consumed across all heritage groups. As a result, its generalizability is limited. Rather, its strength lies in capturing the foods most strongly linked to established dietary quality indices. This limitation is consistent with a broader literature showing that dietary intake, index adherence, and index performance all vary substantially by Hispanic/Latino heritage and acculturation.

Research using data from the Hispanic Community Health Study/Study of Latinos (HCHS/SOL) has shown that dietary quality scores are highest among Mexican heritage and lower among Puerto Rican heritage adults (42), and greater acculturation is associated with declining diet quality driven by lower plant-food intake (43). No single dietary index, whether that is based the HEI, AHEI, Dietary Approaches to Stop Hypertension (DASH), the Mediterranean Diet (MED), Alternative Mediterranean Diet (AMED) scores, has shown superior cardiovascular risk stratification in Hispanic/Latino adults; rather, index performance itself varies by Hispanic/Latino heritage (44, 45). Together, this underscores that heritage and acculturation-specific calibration, more than dietary index selection, is critical to capturing dietary quality and cardiovascular risk in this population, a rationale that motivates SABE’s continued development.

This study has strengths and limitations. SABE was adapted from a screener reflecting foods considered optimal under HEI-2020, with commonly reported NHANES examples, and its scoring algorithm showed adequate alignment with HEI scores replicated across two independent NHANES samples, supportive the representativeness of its items. However, SABE may not fully capture all Hispanic/Latino diets given the NHANES cohort’s predominantly Mexican American composition. AHEI scoring may also underrepresent cardiovascular risk in this population because NHANES lacks trans-fat data, an important AHEI subcomponent. Because our estimates relied on simulated screener responses inferred from dietary recall data rather than direct participant responses, they may not fully represent actual intake and could be optimistically biased. Validating the screener using data collected with the SABE tool administered directly and concurrently to gold standard dietary assessments, alongside validation of our scoring algorithm in Hispanic/Latino adults of diverse backgrounds, would address these limitations and further establish its utility in clinical, community and research settings.

## CONCLUSION

SABE is a brief, feasible dietary screener with strong validity and reproducibility for assessing diet quality in Hispanic/Latino adults. Using nationally representative NHANES dietary recall data, the SABE scoring algorithm demonstrated strong correlation with HEI-2020 scores and moderate correlation with AHEI- 2010 scores, a finding consistent with evidence that index performance itself may vary by Hispanic/Latino heritage group. These findings suggest that SABE captures meaningful variation in overall diet quality as measured by established dietary indices while remaining practical for use in time-constrained settings. SABE addresses an important gap in clinical practice and public health research. Given the limited availability of culturally and linguistically adapted dietary screeners for this population, SABE addresses an important gap in clinical practice and public health research. Future iterations of SABE will incorporate culturally adapted items probing lard use (yes/no/don’t know), distinguish tortilla types (refined flour tortillas and tortilla chips versus whole corn tortillas), and reclassify fruit juice within the sugar-sweetened beverage item rather than the whole-fruit category. Future work will focus on data collection in which the SABE screener is administered directly, alongside gold standard dietary assessment tools, and in Hispanic/Latino adults of diverse heritage— steps that will be essential to confirm SABE’s real-world utility.

## Data Availability

All data produced in the present study are available upon reasonable request to the authors

https://github.com/VasuShandar/SABE_Screener_Scoring_Manuscript

## Author Contributions

WC, GDW and RQ developed the Penn Healthy Diet, the English scale from which SABE was adapted. MSP and CWC conceptualized and designed the study. VS and RQ performed quantitative data analysis. MSP and VM wrote the first draft. All authors (MSP, VS, RQ, VM, KTC, GDW, CC) reviewed and commented on subsequent drafts of the manuscript. MSP and GW obtained funding resources.

We thank Julia Lewandowski for administrative support in the conduct of the SABE study.

## Abbreviations

AHEI: Alternative Healthy Eating Index
AMED: Alternative Mediterranean Diet
CI: Confidence intervals
DASH: Dietary Approaches to Stop Hypertension
FPED: Food Patterns Equivalents Database
HCHS/SOL: Hispanic Community Health Study/Study of Latinos
HEI: Healthy Eating Index
I-CVI: item-level content validity index
MED: Mediterranean Diet
NHANES: National Health and Nutrition Examination Survey
PHD: Penn Healthy Diet
SABE: Screener of Alimentary Behaviors and Eating

