## Supplemental Tables 1-5 for "Development and Scoring of a Diet Quality Screener for Hispanic/Latino Adults Using National Health and Nutrition Examination Survey Data: The ¿SABE Lo Que Come? Tool"

**Co-Authors:**

**Margaret S. Pichardo, MD, PhD, MPH**, Resident Physician, Department of Surgery, University of Pennsylvania, Philadelphia, PA, USA, 19104

**Vasu Shandar**, **BS**, Medical Student, Perelman School of Medicine, University of Pennsylvania, Philadelphia, PA 19104 |

**Ryan Quinn**, MPH, Biostatistician, School of Nursing, University of Pennsylvania, Philadelphia, PA 19104 |

**Victoria McReynolds**, **BS**, Medical Student, Perelman School of Medicine, University of Pennsylvania, Philadelphia, PA 19104 |

**Kate Townsend Creasy, PhD**, Assistant Professor, Department of Biobehavioral Health Sciences, School of Nursing, Division of Translational Medicine and Human Genetics, Perelman School of Medicine, University of Pennsylvania, Philadelphia, PA 19104 |

**Gary D. Wu, MD**, Professor, Division of Gastroenterology and Hepatology, Perelman School of Medicine, University of Pennsylvania, Philadelphia, PA 19104 |

**Charlene Compher**, **PhD, RD, FASPEN**, Emeritus Professor, School of Nursing, University of Pennsylvania, Philadelphia, PA 19104 |

**Corresponding Author:** Margaret S. Pichardo, MD, PhD, MPH | Department of Surgery, University of Pennsylvania, 3400 Spruce Street, Maloney 4, Philadelphia, PA, 19104 | Email:  | Phone: (267) 581-3910

**Supplementary Table 1. Percentage (of total maximum score) intake of HEI and AHEI components by Hispanic ethnicities compared to non-Hispanic adults.**

| **Component** | **Non-Hispanic** | **Mexican American** | | **Other Hispanic** |
| --- | --- | --- | --- | --- |
| ***HEI-2020 COMPONENTS*** | ***Percentage (%) of Maximum Score*** | | | |
| **Total HEI Score** | 49.4 | 49.9 | | 51.9 |
| **Total Vegetables** | 58.8 | 62.9 | | 66.0 |
| **Greens & Beans** | 29.0 | 37.9 | | 46.6 |
| **Total Fruit** | 35.0 | 45.7 | | 46.1 |
| **Whole Fruit** | 38.1 | 47.8 | | 46.7 |
| **Whole Grain** | 24.0 | 18.7 | | 19.0 |
| **Total Dairy** | 48.1 | 44.2 | | 42.9 |
| **Total Protein** | 84.2 | 86.3 | | 83.5 |
| **Seafood/Plant Protein** | 46.5 | 51.7 | | 49.7 |
| **Fatty Acid Ratio** | 48.5 | 52.2 | | 51.7 |
| **Sodium*** | 44.5 | 42.2 | | 46.2 |
| **Refined Grain*** | 63.0 | 46.0 | | 53.4 |
| **Saturated Fat*** | 52.0 | 60.1 | | 64.0 |
| **Added Sugar*** | 68.3 | 70.0 | | 72.6 |
| ***AHEI-2010 COMPONENTS*** | ***Percentage (%) of Maximum Score*** | | | |
| **Total AHEI Score** | 38.9 | 38.2 | 38.7 | |
| **Vegetables (not potatoes)** | 39.1 | 41.6 | 39.5 | |
| **Whole Fruit** | 18.1 | 24.4 | 24.4 | |
| **Whole Grains** | 16.0 | 11.8 | 11.5 | |
| **Sugar Sweetened Beverages & Juice*** | 52.8 | 35.3 | 41.7 | |
| **Nuts & Legumes** | 39.5 | 45.9 | 43.2 | |
| **Red/Processed Meat*** | 52.4 | 53.9 | 55.9 | |
| **Omega-3 (EPA+DHA)** | 21.2 | 22.5 | 23.1 | |
| **PUFA (% of energy)** | 71.0 | 69.2 | 63.6 | |

* Denotes component is reverse scored

**Supplemental Table 2. Retained scoring items in Hispanic/Latino Screener relative to Penn Healthy Diet Screener.**

Items with a total HEI-2020 score above |ρ| ≥ 0.3 threshold were retained.

| **Screener Item** | **Direction** | **Highest Correlation Coefficient (ρ*)*** | **Associated HEI Category** |
| --- | --- | --- | --- |
| **ITEMS INCLUDED IN SABE SCORING ALGORITHM** | | | |
| **Fruit Juice** | Forward | *0.45* | *Total Fruit* |
| **Whole Fruit** | Forward | *0.78* | *Whole Fruit* |
| **Dark Green Veg** | Forward | *0.53* | *Greens & Beans* |
| **Whole Grains** | Forward | *0.99* | *Whole Grain* |
| **Milk** | Forward | *0.52* | *Total Dairy* |
| **Nuts/Seeds** | Forward | *0.40* | *Seafood/Plant Protein* |
| **Seafood** | Forward | *0.42* | *Seafood/Plant Protein* |
| **Plant Protein** | Forward | *0.72* | *Greens & Beans* |
| Red/Orange Veg | Forward | *0.30* | *Total Veg* |
| **Refined Grains** | Reverse | *–0.66* | *Refined Grain* |
| **Cheese** | Reverse | *–0.49* | *Saturated Fats* |
| **Sweet Beverages** | Reverse | *–0.54* | *Added Sugar* |
| **ITEMS EXCLUDED FROM SABE SCORING ALGORITHM BUT INCLUDED IN PHD (below \|ρ\| ≥ 0.3 threshold)** | | | |
| Olive/Veg Oil | — | *0.19* | *Total Veg* |
| Full-Fat Dairy | — | *0.24* | *Total Dairy* |
| Butter/Gravy | — | *–0.15* | *Fatty Acid* |
| **ITEMS NOT IN SABE OR PHD SCORING ALGORITHM** | | | |
| Yogurt | — | *0.22* | *Whole Fruit* |
| Eggs | — | *ρ = 0.16* | *Total Protein* |
| Poultry | — | *ρ = 0.24* | *Total Protein* |
| Red Meat | — | *ρ = –0.21* | *Fatty Acid* |
| Cured Meat | — | *ρ = –0.19* | *Sodium* |
| Desserts | — | *ρ = –0.19* | *Saturated Fats* |

**Supplemental Table 3. Regression weights predicting total HEI-2020 from all 24 screener items, NHANES 2017–18**.

All 18 continuous and 6 yes/no items included. Items ordered by absolute standardized β within each section.

| **Screener Item** | **β** | **Standardized β** | **p-value** |
| --- | --- | --- | --- |
| **FORWARD SCORED — 9 items (higher intake = more points)** | | | |
| *Plant Protein* | +2.79 | 0.35 | <.0001 |
| *Whole Fruit* | +3.50 | 0.27 | <.0001 |
| *Whole Grains* | +2.74 | 0.24 | <.0001 |
| *Nuts/Seeds* | +2.18 | 0.18 | <.0001 |
| *Fruit Juice* | +2.55 | 0.15 | <.0001 |
| *Dark Green Veg* | +2.65 | 0.11 | <.0001 |
| *Milk* | +1.25 | 0.08 | 0.0001 |
| *Seafood* | +1.26 | 0.07 | 0.0001 |
| *Red/Orange Veg* | +1.00 | 0.04 | *0.0317* |
| **REVERSE SCORED — 3 items (lower intake = more points)** | | | |
| *Refined Grains* | –2.62 | –0.24 | <.0001 |
| *Cheese* | –2.07 | –0.16 | <.0001 |
| *Sweet Beverages* | –0.81 | –0.08 | <.0001 |
| **NOT IN SIMPLE SCORING ALGORITHM — continuous items** | | | |
| *Desserts* | –1.44 | –0.13 | <.0001 |
| *Yogurt* | +3.77 | 0.09 | <.0001 |
| *Red Meat* | –1.29 | –0.09 | <.0001 |
| *Cured Meat* | –1.62 | –0.08 | <.0001 |
| *Eggs* | +0.51 | 0.04 | 0.0161 |
| *Poultry* | +0.40 | 0.03 | *0.1007* |
| **YES/NO BEHAVIORAL ITEMS** | | | |
| *Butter/Gravy* | –5.13 | –0.09 | <.0001 |
| *Olive/Veg Oil* | +2.73 | 0.06 | 0.0006 |
| *Full-Fat Dairy* | –2.43 | –0.06 | 0.0029 |
| *Sugar/Honey* | +0.43 | 0.02 | *0.5041* |
| *Cream in Coffee* | –0.52 | –0.02 | *0.4882* |
| *Artificial Sweet* | +0.11 | 0.00 | *0.9098* |
| **INTERCEPT** | **53.77** | **R² = 0.649** | |

**Supplementary Table 4**. **Regression weights predicting total AHEI-2010 from all 24 screener items, NHANES 2017–18.**

All 18 continuous and 6 yes/no items included. Items ordered by absolute standardized β within each direction.

| **Screener Item** | **β** | **Standardized β** | **p-value** |
| --- | --- | --- | --- |
| **POSITIVE β — higher intake predicts higher AHEI** | | | |
| *Whole Fruit* | +3.12 | 0.27 | <.0001 |
| *Plant Protein* | +1.76 | 0.25 | <.0001 |
| *Nuts/Seeds* | +2.35 | 0.22 | <.0001 |
| *Seafood* | +2.45 | 0.16 | <.0001 |
| *Whole Grains* | +1.43 | 0.14 | <.0001 |
| *Red/Orange Veg* | + 2.79 | 0.13 | <.0001 |
| *Eggs* | +1.32 | 0.13 | <.0001 |
| *Dark Green Veg* | +2.20 | 0.10 | <.0001 |
| *Olive/Veg Oil (Y/N)* | +1.68 | 0.05 | 0.0070 |
| *Cream in Coffee (Y/N)* | +1.36 | 0.05 | 0.0197 |
| **NEGATIVE β — higher intake predicts lower AHEI** | | | |
| *Sweet Beverages* | –2.99 | –0.33 | <.0001 |
| *Red Meat* | –3.21 | –0.27 | <.0001 |
| *Cured Meat* | –4.08 | –0.23 | <.0001 |
| *Fruit Juice* | –1.66 | –0.11 | <.0001 |
| *Cheese* | –0.48 | –0.04 | 0.0158 |
| *Sugar/Honey (Y/N)* | –0.76 | –0.03 | *0.1352* |
| *Milk* | –0.36 | –0.03 | *0.1505* |
| *Desserts* | –0.26 | –0.03 | *0.1241* |
| *Refined Grains* | –0.23 | –0.02 | *0.1765* |
| *Butter/Gravy (Y/N)* | –0.35 | –0.01 | *0.6819* |
| *Full-Fat Dairy (Y/N)* | –0.31 | –0.01 | *0.6315* |
| *Yogurt* | –0.22 | –0.01 | *0.7388* |
| *Poultry* | –0.17 | –0.01 | *0.3804* |
| *Artificial Sweet (Y/N)* | +0.14 | 0.00 | *0.8549* |
| **INTERCEPT** | **38.74** | **R² = 0.724** | |

**Supplemental Table 5. Spearman correlations between simulated SABE screener items and AHEI-2010 components in Hispanic/Latino adults, NHANES 2017–18 (N = 1,126).**

| **Screener Item** | **Direction** | **Highest Correlation Coefficient (ρ*)*** | **Associated AHEI Category** |
| --- | --- | --- | --- |
| **ITEMS INCLUDED IN SABE SCORING ALGORITHM** | | | |
| **Fruit Juice** | Forward | –0.39 | SSB + Juice |
| **Whole Fruit** | Forward | 0.99 | Whole Fruit |
| **Dark Green Veg** | Forward | 0.44 | Vegetables |
| **Whole Grains** | Forward | 0.99 | Whole Grain |
| **Milk** | Forward | –0.13 | Sodium |
| **Nuts/Seeds** | Forward | 0.51 | Nuts/Legumes |
| **Seafood** | Forward | 0.58 | Omega-3 |
| **Plant Protein** | Forward | 0.72 | Nuts/Legumes |
| Red/Orange Veg | Forward | 0.55 | Vegetables |
| **Refined Grains** | Reverse | –0.50 | Sodium |
| **Cheese** | Reverse | –0.42 | Sodium |
| **Sweet Beverages** | Reverse | –0.83 | SSB + Juice |
| **ITEMS EXCLUDED FROM SABE SCORING ALGORITHM BUT INCLUDED IN PHD (below \|ρ\| ≥ 0.3 threshold)** | | | |
| Olive/Veg Oil | — | 0.27 | PUFA |
| Full-Fat Dairy | — | 0.12 | Whole Grain |
| Butter/Gravy | — | 0.02 | SSB + Juice |
| **ITEMS NOT IN SABE OR PHD SCORING ALGORITHM** | | | |
| Yogurt | — | 0.19 | Whole Fruit |
| Eggs | — | 0.57 | Omega-3 |
| Poultry | — | 0.22 | Red/Proc Meat |
| Red Meat | — | –0.64 | Red/Proc Meat |
| Cured Meat | — | –0.56 | Red/Proc Meat |
| Desserts | — | –0.16 | Sodium |
